# The demography and apparent risk factors of COVID-19-related suicides in Bangladesh in a seven-month period of the pandemic

**DOI:** 10.1101/2020.08.11.20171272

**Authors:** Sadia Noosrat Boshra, Mohammad Mohiminul Islam, Mark D. Griffiths

## Abstract

In addition to physical health, the COVID-19 pandemic has significantly affected the global economy and mental health. The economic and social impacts have initiated many psychological consequences for individuals. In severe cases, these consequences have led to suicidal behavior among individuals as was witnessed in previous epidemics and pandemics. A few previous studies have been published with regard to COVID-19 suicides in Bangladesh. However, all them concerned more unusual cases of suicide rather than a comprehensive overview of suicides in Bangladesh during the pandemic. Therefore, the present study examined all COVID-19 suicide cases from March 1, 2020, to September 30, 2020, as reported in Bangladeshi online media news portals. A total of 37 suicide cases were identified (65% male; 65% married) with hanging being the most common method used (73%). Just under half the suicides were economically-related reasons due to lock-down-related unemployment (45.9%). These results shed light on the topic with a clearer understanding of the apparent causalities influencing individuals to suicide. Furthermore, it may help Bangladeshi policymakers to take necessary action to prevent further suicides.

## 1. Introduction

The coronavirus disease-2019 (COVID-19) global pandemic has brought about a dramatic change to individuals’ occupational and social behavior including social distancing, quarantining, staying in isolation, and working from home. The pandemic has severely disrupted the global economy and has resulted in unemployment and poverty for millions of individuals (Kawohl & Nordt, 2020). In addition to economic and physical health impact, the pandemic has impacted the mental health of many. In the most severe cases, these consequences have led to suicidal behavior among individuals as was witnessed in the 1918-1919 flu pandemic and in the 2003 severe acute respiratory syndrome (SARS) epidemic (Gunnell et al., 2020). Consequently, it is imperative to monitor the general population’s suicidal behavior in response to the COVID-19 pandemic.

Previously, a few COVID-19 suicide cases were reported in Bangladesh (Bhuiyan et al., 2020; Mamun & Griffiths, 2020; Mamun et al., 2020a; Mamun, Bodrud-Doza, et al., 2020). However, these all concerned more unusual cases of suicide rather than a comprehensive overview of suicides in Bangladesh during the pandemic. Therefore, the present study attempted to document all the COVID-19-related suicide cases published in Bangladeshi media reports to better understand the apparent risk factors and risk groups in order to aid prevention of future suicidal instances.

## 2. Methods

Given that there is no national suicide surveillance system or database available in Bangladesh, online search engines (*Google* and *Bing*) were used to collect retrospective cases of COVID-19-related suicide published in Bangladeshi online media news portals between March 1, 2020, and September 30, 2020. Similar methods of data collection have been used previously in Bangladeshi studies (e.g., Mamun et al., 2020b; Arafat et al., 2018). Although the first case of COVID-19 in the country was reported on March 8, 2020 (Siam et al., 2020), the search was performed from March 1, 2020 because of the prevalent concern and suspicion about COVID-19 before the first confirmed case (NEWAGE Bangladesh, 2020). The keywords used for search were the Bangla language words for suicide (‘Attohottma’), coronavirus in Bangladesh (‘Bangladeshe corona’), COVID-19 in Bangladesh (‘Bangladeshe COVID-19’), and ‘lockdown’. Two of the authors searched independently and then the search results were combined. Duplications were removed and suicide cases that related to COVID-19 comprised the dataset.

## 3. Results

The search identified 37 confirmed COVID-19-related suicide cases reported by Bangladeshi press media (24 males and 13 females). The first instance of COVID-19-related suicide occurred on March 23, 2020. The mean age of the suicides was 35.2 years with a range of 10 to 58 years. Most of the suicides occurred among those who were married (64.9%) and lived in a rural setting (75.7%). The principal method of committing suicide was hanging (n=27), followed by consuming poison/pesticide (n=5), jumping from a height (n=4), and setting oneself on fire (n=1). In most of the cases, multiple factors appeared to contribute to the suicide. In some cases, one risk factor appeared to lead to another (e.g., unemployment leading to family arguments). The main reasons underlying the suicides appeared to be lockdown-related unemployment, depression, family arguments, fear, hunger, poverty, debt, parental abuse, xenophobia, social stigma, parental dispute, and lack of access to treatment. In relation to occupation, one individual was a politician holding a public office, two were government service holders, eight did not report any profession, three were students, and the remaining 23 cases were non-governmental professionals. The majority of the cases or a close family member (n=17) lost their job due to COVID-19 or were day-earners and unable to go to work due to COVID-19-related lock-down. A number of cases were also reported where the victim committed suicide because of failing to pay loan installments to non-governmental organizations (NGOs) and subsequent pressure and/or humiliation from NGO representatives. Only four of the suicides were reported as having COVID-19. One recovered from COVID-19 before committing suicide and the remaining cases did not report whether the individuals had a COVID-19 diagnosis or not. Only one case had a history of mental health issue such as a previous suicide attempt. The details of all 37 cases are presented in Table 1.

**Table 1.** Distribution of demographics, suicide, and reporting variables of the incidents (N=37).

| Demographic variable | Frequency | Percentage |
| --- | --- | --- |
| *Gender* |  |  |
| Male | 24 | 64.9 |
| Female | 13 | 35.1 |
| *Age (years)* |  |  |
| 10-19 | 4 | 10.8 |
| 20-29 | 6 | 16.2 |
| 30-39 | 12 | 32.4 |
| 40-49 | 9 | 24.3 |
| 50-59 | 5 | 13.5 |
| Unknown | 1 | 2.7 |
| *Citizenship* |  |  |
| Bangladeshi | 36 | 97.3 |
| Foreign citizen | 1 | 2.7 |
| *Occupation type* |  |  |
| Non-governmental | 18 | 48.7 |
| Homemaker | 5 | 13.5 |
| Governmental | 2 | 5.4 |

| Student | 3 | 8.1 |
| --- | --- | --- |
| Politician | 1 | 2.7 |
| Not reported | 8 | 21.6 |
| *Marital status* |  |  |
| Married | 24 | 64.9 |
| Unmarried | 1 | 2.7 |
| Widowed | 1 | 2.7 |
| Divorced | 1 | 2.7 |
| Not reported | 7 | 18.9 |
| Minor (<18 years) | 3 | 8.1 |
| *Residence* |  |  |
| Rural | 28 | 75.7 |
| Urban (district/port cities) | 9 | 24.3 |
| Suicide variable | Frequency | Percentage |
| *Method of suicide* |  |  |
| Hanging | 27 | 73.0 |
| Consumption of poison/pesticide | 5 | 13.5 |
| Falling from a height | 4 | 10.8 |
| Setting oneself on fire | 1 | 2.7 |
| *Apparent reason for suicide* |  |  |
| Lockdown related unemployment | 17 | 45.9 |
| Depression | 13 | 35.1 |
| Family arguments | 8 | 21.6 |
| Fear | 6 | 16.2 |
| Debt | 6 | 16.2 |
| Hunger | 4 | 10.8 |
| Poverty | 3 | 8.1 |
| Parental abuse | 2 | 5.4 |
| Xenophobia | 1 | 2.7 |
| Social stigma | 1 | 2.7 |
| Parental dispute | 1 | 2.7 |
| Lack of access to treatment | 1 | 2.7 |
| *COVID-19 diagnosis status* |  |  |
| Positive | 4 | 10.8 |
| Recovered | 1 | 2.7 |
| Not reported | 32 | 86.5 |

## 4. Discussion

The present study examined different variables (i.e., gender, age, occupation, COVID-19 diagnosis, marital status, residence, method of suicide, and apparent suicide reasons) in order to better understand the determinants associated with COVID-19-related suicides in Bangladesh. The findings support the previous Bangladeshi findings in relation to apparent reasons for the suicide (Bhuiyan et al., 2020; Mamun & Griffiths, 2020; Mamun et al., 2020a). However, given that the pandemic is unprecedented, and this is the first comprehensive study to document all COVID-19-related suicides in Bangladeshi setting based on available evidence in a given period of time, no comparison can be made to any previous study in terms of suicidal risk factors and vulnerable groups. Therefore, the present study provides valuable information about factors are associated with suicide during the pandemic.

The first case of COVID-19-related suicide due to fear occurred on March 23, 2020, in contrast to the case reported previously by Mamun and Griffiths (2020) on March 25. This may simply have been due to the different search engines or strategies used. The highest number of suicides occurred during June 2020. This coincided with the highest number of COVID-19 cases being reported in Bangladesh in the same month (see table 2). The present study also found that males were more likely than females to commit suicide due to COVID-related reasons. Although suicide is more common in males compared to females globally, Arafat (2019) reported the opposite in Bangladesh both in the ratio of actual suicides and suicidal attempts. Therefore, it is noteworthy that the incidence of COVID-19-related suicides was higher among males than females. This is most likely because males are the principal wage earners for their families in Bangladeshi society and the economic impact affected males far greater than females.

**Table 2.**
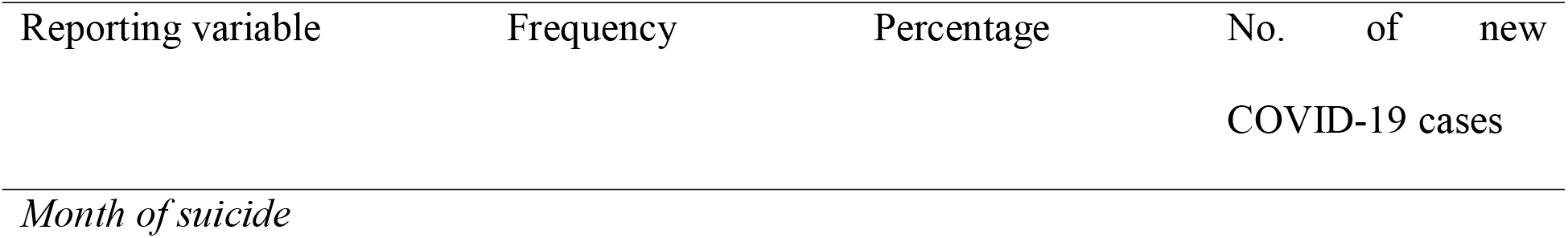
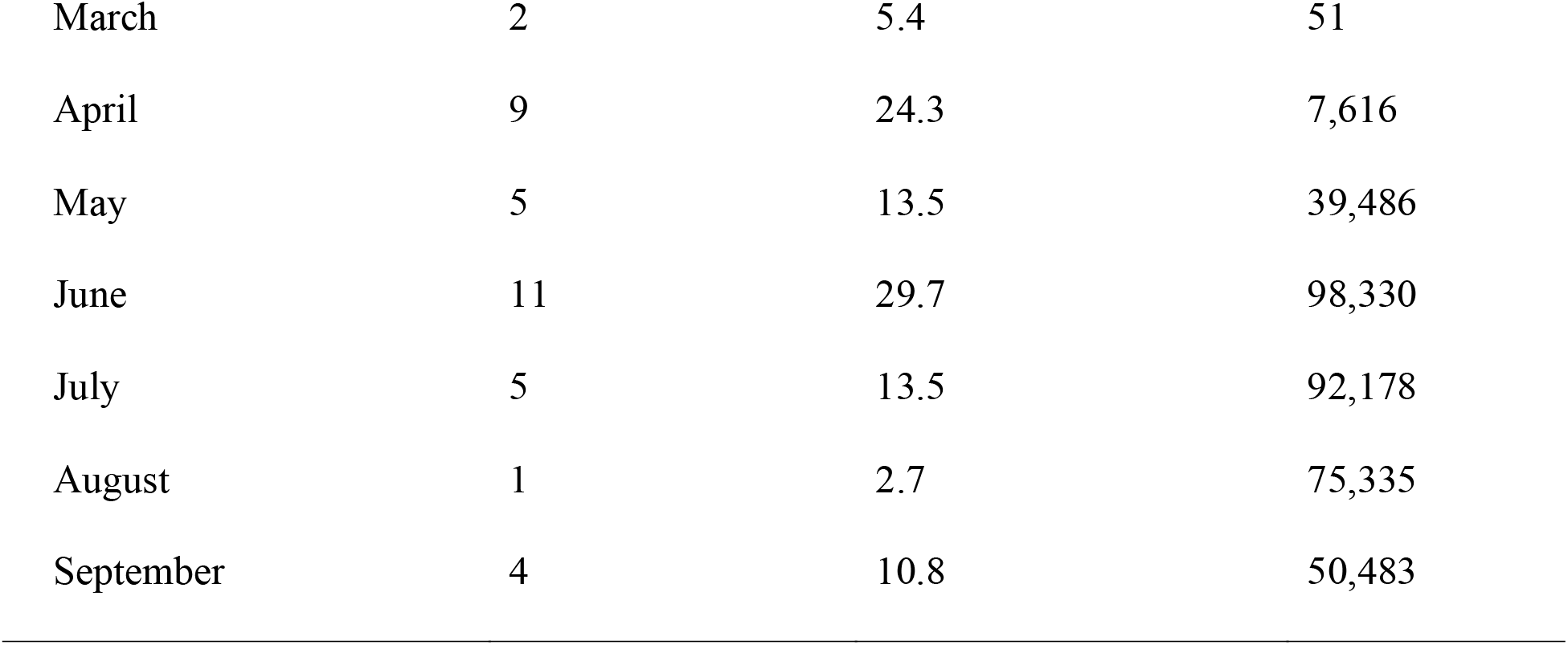
Reporting variable of suicide incidents (N=37) and new COVID-19 cases.

| Reporting variable | Frequency | Percentage | No. of new<br>COVID-19 cases |
| --- | --- | --- | --- |
| *Month of suicide* |  |  |  |
| March | 2 | 5.4 | 51 |
| April | 9 | 24.3 | 7,616 |
| May | 5 | 13.5 | 39,486 |
| June | 11 | 29.7 | 98,330 |
| July | 5 | 13.5 | 92,178 |
| August | 1 | 2.7 | 75,335 |
| September | 4 | 10.8 | 50,483 |

Unsurprisingly, COVID-19-related unemployment was the primary apparent suicide reason among other reasons reported. Apart from fear of COVID-19 and xenophobia, all other apparent reasons for suicide were somewhat related to the individual’s economic status and lack of money being earned. Like many other countries, the Bangladeshi government implemented a nationwide lockdown two weeks after the first confirmed case. The 66-day lockdown significantly impacted low-income families, particularly those who live on a day-to-day income with no savings. During the lockdown, offices were closed, public transportation was shut, and gatherings were suspended (Siam et al., 2020). Consequently, many businesses temporarily or permanently closed, many individuals unemployed. The economic impact was sustained even after the national lockdown ended as indicated by the suicides reported in August and September 2020 due to the economic crisis. Given that, the government employees were economically protected, the number of suicides in this group (n=2) was much lower than others, and one case was because of fear of COVID-19 and depression.

The present study also found that the majority of the suicide cases comprised individuals that were not diagnosed with COVID-19. This indicates that other factors were more salient in determining the apparent reasons for suicide. Hanging was the most common form of suicide, similar to previous studies among the Bangladeshi population (e.g., Arafat et al., 2018). The higher incidence of suicides among those who were married and lived in rural settings has also been found previously among the Bangladeshi population (Shah et al., 2017). However, it should be noted that that data in the present study comprised published press reports identified utilizing online search engines. Therefore, readers should exercise caution in interpreting the results and not to generalize to the entire population because of the associated limitations of data collection utilizing media reports. This method is likely to have led to an under-reporting of suicides given that not all suicides will have been reported in the media and that some deaths may not have been as suicides.

## 5. Conclusion

The present study identified that COVID-19-related unemployment appeared to be the principal cause of suicide based on the data collected. Males were more vulnerable than females to suicide. Very few of the suicides were diagnosed with COVID-19 and was much less as a possible risk factor than economically-related reasons. While the study found that only one suicide was reported with an individual having a pre-existing mental health issue, there is always the possibility that other individuals had pre-existing mental health issues but were not made public by the families and/or the media outlet. Finally, the findings of the present study may help the country’s policymakers to extend and implement initiatives to protect high-risk groups and minimize risk factors. Additionally, the study will also help researchers in the field to do comparative analysis of suicides before, during and after the pandemic.

## Data Availability

Data is available upon request.

## Informed consent

Informed consent is non-applicable to this work.

## Author’s contribution

Conceptualization: MMI; data collection and analysis: SNB and MMI; manuscript draft writing: SNB and MMI; rewriting, editing and finalization of the manuscript: MDG.

## Acknowledgements

None

## References

Arafat S. M. Y. (2019). Females are dying more than males by suicide in Bangladesh. Asian Journal of Psychiatry, 40, 124–125. https://doi.org/10.1016/j.ajp.2018.10.014

Arafat, S. M. Y., Mali, B., & Akter, H. (2018). Demography and risk factors of suicidal behavior in Bangladesh: A retrospective online news content analysis. Asian Journal of Psychiatry, 36, 96–99. https://doi.org/10.1016/j.ajp.2018.07.008

Bhuiyan, A. K. M. I., Sakib, N., Pakpour, A. H., Griffiths, M. D., & Mamun, M. A. (2020). COVID-19-Related suicides in Bangladesh due to lockdown and economic factors: Case study evidence from media reports. International Journal of Mental Health and Addiction, 1–6. Advance online publication. https://doi.org/10.1007/s11469-020-00307-y

Government of Bangladesh (2020). Coronavirus disease 2019 (COVID-19) information. Retrieved October 11, 2020, from: https://corona.gov.bd/

Gunnell, D., Appleby, L., Arensman, E., Hawton, K., John, A., Kapur, N., Khan, M., O’Connor, R. C., Pirkis, J., & COVID-19 Suicide Prevention Research Collaboration (2020). Suicide risk and prevention during the COVID-19 pandemic. The Lancet Psychiatry, 7(6), 468–471. https://doi.org/10.1016/S2215-0366(20)30171-1

Kawohl, W., & Nordt, C. (2020). COVID-19, unemployment, and suicide. The Lancet Psychiatry, 7(5), 389–390. https://doi.org/10.1016/S2215-0366(20)30141-3

Mamun, M. A., Bodrud-Doza, M., & Griffiths, M. D. (2020). Hospital suicide due to non-treatment by healthcare staff fearing COVID-19 infection in Bangladesh? Asian Journal of Psychiatry, 54, 102295. Advance online publication. https://doi.org/10.1016/j.ajp.2020.102295

Mamun, M. A., Chandrima, R. M., & Griffiths, M. D. (2020a). Mother and son suicide pact due to COVID-19-related online learning issues in Bangladesh: An unusual case report. International Journal of Mental Health and Addiction. Advance online publication. https://doi.org/10.1007/s11469-020-00362-5

Mamun, M. A., & Griffiths, M. D. (2020). First COVID-19 suicide case in Bangladesh due to fear of COVID-19 and xenophobia: Possible suicide prevention strategies. Asian Journal of Psychiatry, 51, 102073. https://doi.org/10.1016/j.ajp.2020.102073

Mamun, M. A., Misti, J. M., & Griffiths, M. D. (2020b). Suicide of Bangladeshi medical students: Risk factor trends based on Bangladeshi press reports. Asian Journal of Psychiatry, 48, 101905. https://doi.org/10.1016/j.ajp.2019.101905

NEWAGE Bangladesh (2020). Coronavirus preparedness still a concern in Bangladesh. Retrived October 11, 2020, from: https://www.newagebd.net/article/99712/coronavirus-preparedness-still-a-concern-in-bangladesh

Shah, M., Ahmed, S., & Arafat, S. (2017). Demography and risk factors of suicide in Bangladesh: A six-month paper content analysis. Psychiatry Journal, 2017, 3047025. https://doi.org/10.1155/2017/3047025

Siam, M. H. B., Hasan, M. M., Raheem, E., Khan, M. H. R., Siddiqee, M. H., & Hossain, M. S. (2020). Insights into the first wave of the COVID-19 pandemic in Bangladesh: Lessons learned from a high-risk country. MedRxiv, 2020.08.05.20168674. https://doi.org/10.1101/2020.08.05.20168674

